# Evaluation of the Performance of Filmarray Blood Culture Identification Panel on Detecting Blood Cultures Containing Activated Carbon Powder

**DOI:** 10.1101/2021.12.05.21267325

**Authors:** Chen Chen, Shang He, Chengbin Wang

## Abstract

**Objective:** The FilmArray Blood Culture Identification (BCID) panel is a rapid microfluidic PCR amplification microbial detection system. Several studies have evaluated its clinical performance on the basis of blood culture bottles containing resins. However, proportion of hospitals in China use bottles with carbon power, which the performance of FilmArray has not been fully investigated. Therefore, this study is conducted to explore the accuracy of the panel using blood culture bottles with carbon power.

**Method:** 147 venous blood cultures containing carbon powder were used to assess the microbial and antibiotic resistance detection ability of the FilmArray panel. Outcomes were compared with results of the clinical combination method and their consistency was analyzed.

**Results:** FilmArray detected single microorganism in 121 samples, multiple microorganism in 9 cases and the consistency rate between the two methods was 90.6%. Among the 150 microorganisms detected, 85.1% (40/47) of *staphylococcus* contained the antibiotic resistant *mecA* gene, 15.3% (9/59) of *Enterobacter* detected the *KPC* gene, 7.7% (1/13) of *Enterococcus* has the *vanA* gene and the consistency with their clinical drug-resistant phenotypes were 93.6%, 86.4% and 100%, respectively.

**Conclusion:** The identification rate of the FilmArray BCID panel using venous blood cultures with activated carbon powder was highly consistent with the outcomes of previous researchers using non-carbon powder blood culture bottles. It is capable of providing rapid and reliable results in the detection of pathogens present in automated blood culture systems.

## Introduction

Bloodstream infection (BSI) is a leading cause of death worldwide. Appropriate and timely antimicrobial treatment could significantly reduce the mortality rate for patients with BSI [1]. Conventional antimicrobial treatment often includes the use of broad-spectrum antibiotics, a strategy used due to the lack of specific identification of the causative infectious agent in most cases[2]. However, rapid identification (ID) of microbial pathogens responsible for BSIs is critical to reduce infection-related mortality, particularly for patients admitted to the intensive care units (ICU). So far, blood culture (BC) is an essential tool for BSI diagnosis, but its time-consuming turn-around-time obstructs the timely treatment of patients. Several new technologies such as MALDI-TOF mass spectrometer have been adapted by microbiology laboratories to shorten the time spend on microbial identification [3, 4], but it relies on the outcome of the subculture on the agar plate, which may take several additional days after the blood culture.

In recent years, number of other new rapid BSI detection platforms, such as molecular assays, digital PCR, PCR/ESI-MS technology *et al*, have emerged [5-8], and each platform has its benefits and limitations[9]. The FilmArray Blood Culture Identification (BCID) panel (BioFire Diagnostics Salt Lake City, UT) is a novel rapid diagnostic method based on multiplex polymerase chain reaction and microfluidic technology to detect common sepsis-related pathogens in an hour, including 19 bacteria and 5 yeasts and three antimicrobial resistance genes, using samples from positive blood culture bottles/broths [10]. Previous studies have evaluated the diagnostic accuracy of the FilmArray BCID panel in comparison to conventional laboratory methods[11-14] on the basis of blood culture bottles or bottles containing resins. Our hospital and many hospitals like us in China, however, mainly use blood culture bottles, especially for the inpatients and the ICU department, containing 32ml of complex media and 8ml of charcoal suspension with an average density of 1.0215 g/mL. Although adding charcoal can bring several benefits such as adsorbing antibiotics in the blood, reducing antibiotics interference, improving positive rate of the blood culture and shortening the turn-around-time, the adsorption is also a double-edged sword. It could affect the extraction of nucleic acid sample and the enzyme activity in the PCR reaction system, which may also disturb the detection of FilmArray BCID panel. To the best of our knowledge, the performance of FilmArray BCID panel on blood culture bottle containing activated carbon has not been properly investigated. Therefore, in this study we tested and verified differences between the culture bottles containing carbon powder detected by the FilmArray BCID panel and a combination of conventional clinical laboratory methods.

## Materials and Methods

The study was conducted in a 3A grade hospital with a bed capacity of 4400 in China. The study protocol was reviewed and approved by the Biomedical Research Ethics Committee of the PLA General Hospital. Samples of this study were collected from the inpatients in the hospital from January 2017 to June 2018, and consents were obtained from the patients whose samples were used as per the existing agreement to the PLA General Hospital Medical Microbiology laboratories.

### Study setting and design

All blood cultures were received as per routine work from all wards and ICU of the hospital covered by the Medical Microbiology Laboratory of the PLA. Per routine processes, 8-10mL blood from the adult patient was injected into the blood culture bottle BACT/ALERT® FA and BACT/ALERT® FN (BioMérieux, Shanghai, CN), respectively, and 3-5ml venous blood from pediatric patient were injected into the special BacT/ALERT® PF blood culture bottles. The blood cultures were then loaded into the automated blood culture continuous monitoring system,BacT-ALERT® 3D 120 (BioMérieux, Shanghai,CN). If no signal was received from a blood culture bottle after seven days, the bottle was retrieved and resulted as ‘no growth after seven days (except for the samples which the doctor asks for extending culture time). If the instrument alarm indicated the bottle had possible positive results, it would then be taken out from the instrument for smear and optical microscopy examination. If there were no bacteria found in the microscope field, the bottle would be loaded into the instrument for more culturing until the positive alarm show up again in next seven day, or it will be reported as “no growth”.

### Clinical laboratory identification of positive cultures

For the positive samples examined by the microscope, cultures were isolated and subcultured on Columbia sheep blood agar plate, China blue agar plate and Sabouraud’s agar plate, and the anaerobic cultures were cultured on CDC anaerobic agar culture plate in the CO_2_ incubator. The special bacteria were cultured on its optimal medium according to its cultural properties. After cultured for 16-24 hours, a single colony of each type of colony on the agar plate was identified by VITEK MS (MALDI-TOF mass spectrometer, BioMérieux). If there was no identification result of the mass spectrum database, the microorganism nucleic acid would be extracted for the identification by the next-generation gene sequencing (NGS). This approach was called the clinical combination method.

### Antimicrobial susceptibility test of microorganisms

For the microorganisms in samples identified as bacteria, its antimicrobial susceptibility detection would be detected by VITEK 2 compact (BioMerieux). AST-GP 67 card was used for gram-positive bacteria, AST-GN09 card for non-fermentative gram-negative bacteria and AST-GN13 card for fermentative gram-negative bacteria. Other separate drug sensitivity tests would be added for special bacteria according to the identification specifications of the laboratory, which were tested by Kirb-Bauer (K-B) or Epsilometer test (E-test). The drug sensitivity of fungi was determined by VITEK 2 AST-YS01 (BioMerieux) fungal drug sensitivity system.

The clinical identification and antimicrobial susceptibility detection were called as the clinical combination method.

### Identification of positive cultures by FlimArray BCID panel

Each positive blood culture was run on a single FilmArray BCID panel within eight hours after becoming positive. 100μL of the contents of the blood culture bottle was aspirated, mixed with sample buffer (provided) and loaded into the FilmArray pouch. The pouch was loaded onto the FilmArray instrument, which was connected to a computer system. At the end of the run, a report was automatically generated showing any detectable organism(s) as well as any antimicrobial resistance gene(s) – *mecA, vanA/B* and KPC. (Fig.1)

### Reference strains in the study

American Type Culture Collection (ATCC) strains were used as external controls and set up by the study. Quality control strains of the BacT/ALERT3D were *Staphylococcus aureus* ATCC 25923, *Escherichia coli* ATCC25922 and *Pseudomonas aeruginosa* ATCC27853. Quality control strains of the VITEK MS were *Staphylococcus aureus* ATCC 25923, *Escherichia coli* ATCC 8739 and *Candida albicans* ATCC90029. The quality control strains of VITEK 2 compact were *Escherichia coli* ATCC25922 for AST-GN13 card, *Staphylococcus aureus* ATCC29213 for AST-GP67 card, *Streptococcus pneumoniae* ATCC49619 for AST-GP68 card and *Pseudomonas aeruginosa* ATCC27853 for AST-GN09 card.

### Statistical analyses

In this study, identification and antimicrobial susceptibility of organisms obtained by the FlimArray BCID panel were compared to those obtained by the combination method i.e., the VITEK MS methods and next-generation sequencing. If the VITEK MS did not identify any microbial, then NGS results were used as the clinical results. Identification that was brought down to the nearest precision possible (genus/species/complex/subspecies level achievable) by FlimArray BCID panel with respect to the results seen from the combination method was labeled as ‘precisely identified’; organisms, listed in the panel, that were missed or misidentified (genus/species/complex/ subspecies level), were labelled as either ‘missed’ or ‘misidentified’. The number of blood cultures, and the number of organisms, were all used to evaluate the performance of FlimArray BCID panel in the statistical analyses. Sensitivity, specificity, positive predictive value and negative predictive value as well as consistency were analyzed. Calculations were done using IBM SPSS Statistics for Windows (Version 22.0; IBM Corp, Armonk, New York, United States).

## Results

### Sample information

A total of 147 venous blood culture samples from inpatients were tested in this study and 105 of which were males and 42 were females. The age range was from 18 to 92 years.

### Outcomes of microbial identification

Five blood culture bottles inoculated with known ATCC organisms were used as external controls on FilmArray. All five external controls yielded the desired identification. Among the 147 samples tested by clinical combination methods, 129 of which contained a single microorganism, 9 samples contained more than one type of microorganisms, confirmed by the VITEK MS, and 9 false positive samples were detected. In comparison, FilmArray BCID panel detected 17 negative samples and 130 positive samples from the 147 cases, in which 121 had a single positive microorganism (Table 1) and 9 had more than one type of microorganism (Table 2). In the 121 positive samples, 1 was incorrectly identified and in 9 samples with multiple microorganisms, 1 was identified as a single microorganism by the clinical combination methods. In addition, 1 sample containing two types of microorganisms was identified as negative by the FilmArray BCID and four samples had negative outcome because the corresponding microorganisms were out of panel’s detection range. Moreoever, nine samples with “no growth” detected by combination method were identified as negative samples by NGS test.

**Table 1.** Performance of FilmArray BICD on blood culture samples with one type of organism

| Filmarray BICD | Identification by combination methods | Precisely identified<br>n (%) | Imprecisely identified<br>n (%) | Missed<br>n (%) |
| --- | --- | --- | --- | --- |
| **Gram-Positive Bacteria** |  | 55(98.2) | 1(1.8) | 0 |
| *Staphylococcus* | *Staphylococcus capitis* | 3 | - | - |
|  | *Staphylococcus haemolyticus* | 3 | - | - |
|  | *Staphylococcus hominis* | 26 | - | - |
|  | *Staphylococcus epidermidis* | 9 | - | - |
| *Staphylococcus aureus* | *Staphylococcus aureus* | 4 | 1 | - |
| *Streptococcus* | *Streptococcus oralis* | 2 | - | - |
| *Streptococcus pyogenes* | *Streptococcus pyogenes* | 1 | - | - |
| *Enterococcus* | *Enterococcus faecium* | 6 | - | - |
|  | *Enterococcus faecalis* | 1 | - | - |
| **Gram-Negative Bacteria** |  | 54(100) | 0 | 0 |
| *Enterobacteriaceae* | *Salmonella* | 1 | - | - |
|  | *Enterobacter aerogenes* | 1 | - | - |
| *Escherichia coli* | *Escherichia coli* | 26 | - | - |
| *Klebsiella pneumoniae* | *Klebsiella pneumoniae* | 16 | - | - |
| *Klebsiella oxytoca* | *Klebsiella oxytoca* | 1 | - | - |
| *Pseudomonas aeruginosa* | *Pseudomonas aeruginosa* | 4 | - | - |
| *Acinetobacter baumannii* | *Acinetobacter baumannii* | 3 | - | - |
| *Serratia marcescens* | *Serratia marcescens* | 1 | - | - |
| *Enterobacter cloacae complex* | *Enterobacter cloacae complex* | 1 | - | - |
| **Yeast** |  | 11(100) | 0 | 0 |
| *Candida albicans* | *Candida albicans* | 2 | - | - |
| *Candida glabrata* | *Candida glabrata* | 3 | - | - |
| *Candida parapsilosis* | *Candida parapsilosis* | 4 | - | - |
| *Candida tropicalis* | *Candida tropicalis* | 2 | - | - |
| **Failed to detect** |  | - | - | 8 |
| Not Detected | *Bacillus cereus* | - | - | 1 |
|  | *Cryptococcus neoformans* | - | - | 1 |
|  | *Morganella morganii* | - | - | 1 |
|  | *Pantoea agglomerans* | - | - | 1 |
|  | *Staphylococcus saccharolyticus* | - | - | 1 |
|  | *Myxobacterium* | - | - | 1 |
|  | *bacterium burgeri* | - | - | 1 |
|  | *Staphylococcus pettenkoferi* | - | - | 1 |
| **False positive blood cultures** |  | 9(100) | - | - |
| Not Detected | No bacteria were cultivated | 9 | - | - |
| **Total** |  | 129(93.5) | 1(0.7) | 8(5.8) |

**Table 2.** Performance of FilmArray BICD on blood culture samples with more than one type of organism

| Filmarray | Identification by combination methods | Precisely identified<br>n (%) | Imprecisely identified<br>n (%) | Missed<br>n (%) |
| --- | --- | --- | --- | --- |
| *Enterococcus Pseudomonas aeruginosa* | *Enterococcus Pseudomonas aeruginosa* | 1 | - | - |
| *Enterococcus Escherichia coli* | *Enterococcus avium Escherichia coli* | 1 | - | - |
| *Enterococcus Escherichia coli* | *Enterococcus faecium Escherichia coli* | 1 | - | - |
| *Enterococcus Escherichia coli* | *Enterococcus casseliflavus Escherichia coli* | 1 | - | - |
| *Enterococcus Acinetobacter baumannii* | *Enterococcus faecium Acinetobacter baumannii* | 1 | - | - |
| *Pseudomonas aeruginosa* | *Pseudomonas aeruginosa Hydrophila/Caviae Aeromonas* | - | 1 | - |
| *Enterococcus Staphylococcus aureus* | *Enterococcus faecalis Staphylococcus aureus Streptococcus dysgalactiae* | - | 1 | - |
| *Enterococcus Klebsiella pneumoniae* | *Enterococcus faecium Klebsiella pneumoniae Escherichia hermannii Hydrophila/Caviae Aeromonas* | - | 1 | - |
| *Enterococcus Candida tropicalis* | *Stenotrophomonas maltophilia Candida tropicalis* | - | 1 | - |
| None | *Micrococcus luteus Micrococcus lylae* | - | - | 1 |
| **Total** |  | 5<br>(50.0%) | 4<br>(40.0%) | 1<br>(10.0%) |

In the 147 samples of this study, 150 strains of microorganisms were identified by the combination method, shown in Table 3, and 135 strains were identified by FilmArray BICD, which was 90.0% of the combination method. However, 15 strains of microorganisms were not identified by the combination method, accounting for 10.0%. There were 64 strains of gram-positive bacteria in the accurate identification, accounting for 92.8% (64/69) of the positive bacteria, 59 strains of gram-negative bacteria, accounting for 86.8% (59/68) of the negative bacteria, and 12 strains of fungi, accounting for 92.3% (12/13) of the total bacteria. The 15 strains not identified by the FilmArray BICD were not included in the identification panel.

**Table 3.** Performance of FilmArray BICD on all the organisms detected by combination methods

| Filmarray BICD | Identification by combination methods | Precisely identified n (%) | Imprecisely identified n (%) | Missed n (%) |
| --- | --- | --- | --- | --- |
| **Gram-Positive Bacteria** |  | 63(92.8) | 1(1.6) | 0 |
| **Staphylococcus** | *Staphylococcus capitis* | 3 | / | / |
|  | *Staphylococcus haemolyticus* | 3 | / | / |
|  | *Staphylococcus hominis* | 26 | / | / |
|  | *Staphylococcus epidermidis* | 9 | / | / |
| **Staphylococcus aureus** | *Staphylococcus aureus* | 5 | 1 | / |
| **Streptococcus** | *Streptococcus oralis* | 2 | / | / |
| **Streptococcus pyogenes** | *Streptococcus pyogenes* | 1 | / | / |
| **Streptococcus agalactiae** | *Streptococcus agalactiae* | 1 | / | / |
| **Enterococcus** | *Enterococcus faecium* | 8 | / | / |
|  | *Enterococcus faecalis* | 2 | / | / |
|  | *Enterococcus casseliflavus* | 1 | / | / |
|  | *Enterococcus avium* | 1 | / | / |
|  | *Enterococcus* | 1 | / | / |
| **Gram-Negative Bacteria** |  | 59(86.8) | / | / |
| **Enterobacteriaceae** | *Salmonella* | 1 | / | / |
|  | *Enterobacter aerogenes* | 1 | / | / |
| **Escherichia coli** | *Escherichia coli* | 28 | / | / |
| **Klebsiella pneumoniae** | *Klebsiella pneumoniae* | 16 | / | / |
| **Klebsiella oxytoca** | *Klebsiella oxytoca* | 1 | / | / |
| **Pseudomonas aeruginosa** | *Pseudomonas aeruginosa* | 6 | / | / |
| **Acinetobacter baumannii** | *Acinetobacter baumannii* | 4 | / | / |
| **Serratia marcescens** | *Serratia marcescens* | 1 | / | / |
| **Enterobacter cloacae complex** | *Enterobacter cloacae complex* | 1 | / | / |
| **Yeast** |  | 12(92.3) | / | / |
| **Candida albicans** | *Candida albicans* | 2 | / | / |
| **Candida glabrata** | *Candida glabrata* | 3 | / | / |
| **Candida parapsilosis** | *Candida parapsilosis* | 4 | / | / |
| **Candida tropicalis** | *Candida tropicalis* | 3 | / | / |
| **Failed to detect** |  | / | / | 15 |
| **Not Detected** | *Bacillus cereus* | / | / | 1 |
|  | *Cryptococcus neoformans* | / | / | 1 |
|  | *Morganella morganii* | / | / | 1 |
|  | *Pantoea agglomerans* | / | / | 1 |
|  | *Staphylococcus saccharolyticus* | / | / | 1 |
|  | *Myxobacterium* | / | / | 1 |
|  | *bacterium burgeri* | / | / | 1 |
|  | *Staphylococcus pettenkoferi* | / | / | 1 |
|  | *Streptococcus dysgalactiae* | / | / | 1 |
|  | *Stenotrophomonas maltophilia* | / | / | 1 |
|  | *Hydrophila/Caviae Aeromonas* | / | / | 2 |
|  | *Escherichia hermannii* | / | / | 1 |
|  | *Micrococcus luteus* | / | / | 1 |
|  | *Micrococcus lylae* | / | / | 1 |
| **Total** |  | 134(89.3) | 1(0.7) | 15(10.0) |

### Antibiotic sensitivity test

FilmArray BCID panel contains three drug-resistant genes, namely *mecA* of *Staphylococcus spp, VanA/VanB* of *Enterococcus spp* and KPC of *Enterobacteriaceae*, therefore only these species had the results of drug resistance gene detection. In 150 strains of microorganism detected by combination methods, 40 strains (85.1%) were detected with *mecA* gene among a total of 47 strains of staphylococcus. Of which 37 cases were consistent with the clinical drug resistance phenotype accounting for 93.6%. 1 strain (7.7%) was detected with *vanA* gene among 13 strains of and this finding was consistent with the clinical drug resistance phenotype, accounting for 100%. In 59 strains of *Enterobacterium* detected, 9 strains contained the KPC gene (15.3%), and a total of 52 strains were consistent with the clinical drug resistance phenotype, accounting for 88.1%. Among the detected *Enterobacteriaceae*, 4 strains of *Pseudomonas aeruginosa* and 4 strains of *Acinetobacter baumannii* showed insensitivity to carbapenems antibiotics without the detection of *KPC* gene in their genome. One strain of *Klebsiella oxytoca*, which did not contain the *KPC* gene, showed sensitivity to carbapenems. (Table 4)

**Table 4.** The distribution of antibiotic resistance marker detected by FilmArray BICD and the drug-resistant phenotype of all the related organisms a: S: sensitive; I: inhibited; R: Resistant.

| Species | Numbers<br>n(%) | Resistant genes (FilmArray) |  |  | Antimicrobial<br>susceptibility |  |  |
| --- | --- | --- | --- | --- | --- | --- | --- |
|  |  | *mecA* | *vanA/vanB* | KPC | S<sup>a</sup> | I | R |
| *Staphylococcus aureus* | 5(10.6%) | 1(100) | / | / | 4 | 0 | 1 |
| *Staphylococcus epidermidis* | 9(19.2%) | 9(100) | / | / | 0 | 0 | 9 |
| *Staphylococcus hominis* | 27(57.5%) | 25(92) | / | / | 4 | 0 | 23 |
| *Staphylococcus cephalis* | 3(6.4%) | 3(66.7) | / | / | 1 | 0 | 2 |
| *Staphylococcus haemolyticus* | 3(6.4%) | 2(100) | / | / | 1 | 0 | 2 |
| **Total** | 47 | 40(93.6) | / | / | 10 | 0 | 37 |
| *Enterococcus* | 13(100) | / | 1(7.7) | / | 12 | 0 | 1 |
| **Total** | 13 | / | 1 (100) | / | 12 | 0 | 1 |
| *Enterobacter aerogenes* | 1(1.7%) | / | / | 1(100) | 0 | 0 | 1 |
| *Salmonella* | 1(1.7%) | / | / | 0(100) | 1 | 0 | 0 |
| *Escherichia coli* | 29(49.2%) | / | / | 1(100) | 28 | 0 | 1 |
| *Klebsiella pneumoniae* | 17(28.8%) | / | / | 7(100) | 10 | 0 | 7 |
| *Klebsiella oxytoca* | 1(1.7%) | / | / | 0(100) | 1 | 0 | 0 |
| *Pseudomonas aeruginosa* | 6(10.2%) | / | / | 0(33.3) | 2 | 1 | 3 |
| *Acinetobacter baumannii* | 4(6.8%) | / | / | 0(0) | 0 | 0 | 4 |
| **Total** | 59 | / | / | 9(86.4) | 42 | 1 | 16 |

### Sensitivity and specificity of FilmArray BCID panel

A total of 147 blood culture samples were detected by the two methods. The combination method detected 138 positive culture samples, 129 positive samples of single microorganism, and 9 cases of more than two kinds of microorganism. FilmArray BCID detected 130 positive samples, including 121 single microorganism samples and 9 cases with multiple microorganism. By using the combination method as the reference, the overall sample sensitivity of FilmaArray was 90.6%, the sensitivity of single microorganism positive sample was 93.8%, the sensitivity of multiple microorganisms was 50.0%, and the overall sensitivity of microbial detection was 90.0%. The specificity and positive predictive value were both 100%. The negative predictive value for overall positive blood cultures, blood cultures containing single microorganism and blood cultures containing more than two microorganisms were 40.9%, 52.9% and 64.3%, respectively. The negative predictive value of microorganism was 37.5% (Table 5).

**Table 5.** Performance characteristics of FilmArray BICD compared to combination methods.

| Parameter | All positive blood cultures (n = 139) | Positive cultures with one type of organism (121/139) | Positive cultures with more than one type of organism (9/139) | All organism by detected (134/159) |
| --- | --- | --- | --- | --- |
| Sensitivity | 90.6%<br>95% CI [84.5-94.9] | 93.8%<br>95% CI [88.1-97.3] | 50.0%<br>95% CI [18.7-81.3] | 90.0%<br>95% CI [84.5-94.9] |
| Specificity | 100%<br>95% CI [66.4-100] | 100%<br>95% CI [66.4-100] | 100%<br>95% CI [66.4-100] | 100%<br>95% CI [66.4-100] |
| Positive predictive value | 100%<br>95% CI [N/A] | 100%<br>95% CI [N/A] | 100%<br>95% CI [N/A] | 100%<br>95% CI [N/A] |
| Negative predictive value | 40.9%<br>95% CI [28.1-51.4] | 52.9%<br>95% CI [36.5-68.8] | 64.3%<br>95% CI [49.2-77.0] | 37.5%<br>95% CI [29.2-53.7] |
| Cohen's Kappa | 0.54 | 0.66 | 0.49 | 0.50 |

Positive blood cultures described are organisms that were on the FilmArray BCID repertoire.

## Discussion

A sound correlation was found between the FilmArray and combination methods for identification of organisms and resistance genes. An overall identification sensitivity of 89.3% was achieved for organisms identifiable from all blood cultures by FilmArray. This was slightly lower than the overall sensitivity of 92.6% reported in a separate study by Mokshanand Fhooblall *et al*. Higher sensitivity of FilmArray was seen in positive cultures with a single organism (94.2% [129/137] organisms detected) than in cultures with multiple organism (55.5% [5/9] detected) in our study. This was in line with the previous work which also found lower sensitivity for cultures that detected more than one organism. However, no organism, listed in the panel but not detected by the FilmArray, was picked up by combination methods, which is different from other studies.

FilmArray BCID panel is a product that can be used to rapidly identify microbes in blood culture. Comparing with the traditional blood culture identification method, this method does not require subculture on the agar plate and identify colonies of bacteria (e.g. Meriere’s VITEK 2 Compact system), instead, it relies on the direct multi-PCR amplification with the blood culture without nucleic acid extraction and so approximately 27.9-29.7 h was saved. Moreover, FilmArray can provide information of several common clinical microbial resistance genes, which is beneficial for treating of sepsis patients. It is helpful for the doctors to make quick decisions, and conducive to the management of antibiotics. Two other molecular diagnostic methods, Luminex’s Verigen® System and GenMark’s ePlex system also offers direct microorganism detection from the blood culture, but GN, GP or FP must be separated before identification and different cards are used. Therefore, the integrated detection system of Filmarray is more convenient and time saving in operation.

In terms of performance, FilmArray BCID panel showed a high consistency in the identification of single microbial infection samples with clinical combination methods in this study. The detection rate (90.6%, n=148) of the positive sample with one organism in the samples was similar to that (88.1%, n=2207) of the study carried out by Hossein Salimnia *et al*. and the detection rate of the sample with multiple organisms was also relatively close. [15] Only one case of *Staphylococcus aureus* was misidentified as *Staphylococcus spp*, failing to give a more accurate judgment. The 17 samples tested negative by the Filmarray have lead to a big drop in detection specifity, the reason was that these microorganisms were not within the scope of identification by the panel, which is a point of future improvement of this product. At this point, Filmarray and similar detection systems only covers limited number of pathogen species. For example, Fil,marray detects 24 pathogens (including fungi) and 3 drug-resistantance genes, Verigen could only identify 22 pathogens (no fungi) and 9 drug-resistantance genes and ePlex can detect 29 pathogens, but no drug-resistant genes. In contrast, MALDI-ToF based VITEK® MS platform is able to identify more than 1,000 pathogens, and VITEK 2 can identify 997 antibiotic resistance sensitive phenotypes. Additionally, 2 strains of Staphylococcus hominis and 1 strain of Staphylococcus cephalic with *mecA* gene detected showed their sensitivity to antibiotics (Table 5), indicating that the genotype and phenotype of these strains were not completely consistent, and so gene detection cannot completely replace phenotype detection.

For samples with multiple microbial infections, 4 cases not identified by the Filmarray because the corresponding bacteria were beyond the scope of the panel. In the other 3 cases, the identifications of the combination method were *Enterococcus spp*, but Filmarray detected multiple microorganism, which may be caused by contamination during the detection process. Because the principle of FilmArray BCID panel is PCR technology, contamination is one of the most common problems[16, 17]. How to avoid contamination is also one of the aspects that need to be future improved in this detection reagent.

In this study, all the samples are from blood cultures containing carbon powder, the accuracy of detection of the Filmarray was good comparing with the current clinical combination method, and this was consistent with the outcomes of previous researchers using non-carbon powder blood culture bottles. It is capable of providing rapid [18-20] and reliable results in the detection of pathogens present in automated blood culture systems[11, 21], and it can be applied to the special patients[22-24] and the sterile body fluids, such as cerebrospinal, joint, pleural and ascitic fluids, bronchoscopy samples and abscesses[25-27]. The panel can not only save the patient cost ($30,000 saved per 100 patients tested) but also improve the ASP [28-30]. In addition, because of its simple operation, it reduces the labor intensity and their experience requirement of the staff[31].

## Data Availability

All data produced in the present study are available upon reasonable request to the authors

## Acknowledgements

We thank the PLA General Hospital Clinical Microbiology Laboratory team for the helping. Thank BioMerieux (Shanghai) for providing the FilmArray BCID panel kits.

